# Complementing Tissue Testing with Plasma mutation Profiling Improves Therapeutic Decision Making for Lung Cancer Patients

**DOI:** 10.1101/2021.08.04.21261589

**Authors:** Yukti Choudhury, Min-Han Tan, Jun Li Shi, Augustine Tee, Kao Chin Ngeow, Jonathan Poh, Ruth Rosalyn Goh, Jamie Mong

**Affiliations:** Lucence Diagnostics Pte. Ltd., Singapore; Institute of Bioengineering and Nanotechnology, Singapore; Department of Respiratory and Critical Care Medicine, Changi General Hospital, Singapore; Faculty of Medicine, Imperial College London

## Abstract

**Background:** Tissue biopsy is an integral part of the diagnostic approach to lung cancer. It is however invasive and associated with limitations of tissue heterogeneity. Liquid biopsies may complement tissue testing by providing additional molecular information and may be particularly helpful in patients from whom obtaining sufficient tissue for genomic profiling is challenging.

**Methods:** Patients with suspected lung cancer (n=71) were prospectively recruited. Blood and diagnostic tissue samples were collected within 48 hrs of each other. Plasma cell-free DNA (cfDNA) testing was done using an ultrasensitive amplicon-based next-generation sequencing (NGS) panel (plasma NGS testing). For cases diagnosed as non-small cell lung carcinoma (NSCLC) via histology or cytology, targeted testing for epidermal growth factor receptor (*EGFR*) mutations was performed using tissue biopsy samples, where available (tissue *EGFR* testing). Concordance of clinically actionable mutations between methods and sample types were assessed.

**Results:** For confirmed NSCLC cases (n = 54), tissue *EGFR* test results were available only for 70.3% (38/54) due to sample inadequacies, compared to blood samples for 98.1% (53/54) cases. Tissue *EGFR* testing identified sensitizing *EGFR* (L858R or exon 19 deletion) mutation in 31.6% (12/38) of cases. Plasma NGS identified clinically actionable mutations in 37.7% (20/53) of cases, including *EGFR* mutations in two cases with no tissue EGFR results, and mutations in *KRAS, BRAF* and *MET*. Overall sensitivity of *EGFR* sensitizing mutation detection by plasma NGS was 75% (9/12), and specificity was 100% (25/25) in patients tested in both tissue *EGFR* and plasma NGS (n=37). In this cohort of patients, tissue *EGFR* testing alone informed clinical decisions in 22.2% (12/54) of cases. Adding plasma NGS to tissue *EGFR* testing increased the detection rate of actionable mutations to 42.6% (23/54), representing a near doubling (1.9-fold increase) of clinically relevant findings. The average turnaround time (TAT) of plasma NGS was shorter than standard tissue testing (10 days vs. 29.9 days, p-value <0.05).

**Conclusions:** In the first-line setting, plasma NGS was highly concordant with tissue *EGFR* testing. Plasma NGS increases the detection of actionable findings with shorter time to results. This study outlines the clinical utility of a complementary plasma mutation profiling in the routine management of lung cancer patients.

## INTRODUCTION

Lung cancer is the most common cause of cancer death worldwide, and non-small cell lung cancer (NSCLC) accounts for 85% of all lung cancers, making NSCLC a major cause of mortality^1^. The 5-year survival rate of lung cancer patients is 18.6% and for late-stage NSCLC the 5-year survival rate stands at 6%^2^. The median age of diagnosis of NSCLC is 70 years of age and about 40% of patients are diagnosed with lung cancer at a late stage^2^. Given the age profile and time-scarce outlook for the average lung cancer patient, it is important to create diagnostic tools that are fast, sensitive and accessible by all patients, in particular those of advanced age or cancer stage.

Major progress has been made in the treatment of advanced NSCLC with the identification of specific driver mutations and the development of targeted therapies^3,4^. Although actionable mutations are found in only a subset of patients, progression-free survival was shown to be significantly increased in patients treated with targeted therapy compared to those treated with chemotherapy^5^. Molecular diagnostic testing combined with molecular targeted agents directed against driver mutations in *EGFR, ALK, ROS1, BRAF, MET, RET* and most recently *KRAS* have significantly improved the outcomes for patients with advanced disease harbouring these alterations^6,7^. The most recent National Comprehensive Cancer Network (NCCN) guideline recommendations (Version 5.2021) for the management of NSCLC now include testing for *EGFR, BRAF, ALK, ROS1, RET, KRAS, MET* exon 14 skipping and *NTRK* in non-squamous lung cancer, as part of broader molecular profiling^8^.

Tissue biopsy is the prevailing gold standard for the diagnosis of NSCLC among patients suspected to have lung cancer, and tumor testing is most commonly used for the determination of guideline-recommended biomarkers. In about 15% to 40% of NSCLC cases, comprehensive molecular testing is not feasible due to insufficient tissue samples^9,10^. In the absence of a comprehensive tissue test, a serial testing approach was shown to be successful in only 5% of patients for all 8 guideline-recommended biomarkers^11^. Sampling a single lesion may not capture the complete genomic landscape due to molecular heterogeneity of tumors^12^. The risk of complications is another concern, rising up to 61% with the use of transthoracic needle biopsy, and the incidence of pneumothorax also increases significantly in older patients with obstructive lung disease^13^. Another challenge is the time required for guideline-complete tissue testing which can result in a substantial number of patients initiating chemotherapy before diagnostic results become available, with 19% of *EGFR* mutation or ALK rearrangement positive patients initiating first-line chemotherapy while awaiting their biomarker test results^10^. Liquid biopsies present an alternative approach to tissue-based diagnostic testing, with the use of plasma cfDNA as the substrate for molecular profiling. Tumor alterations identified through routine analysis of clinical tissue samples are detected in cfDNA with a sensitivity of ∼80–90%^14^. Detection sensitivity is influenced by both anatomical sites of disease and tumor burden which in turn correlates with overall circulating tumor DNA (ctDNA) burden^15–17^. A recent study focused on the use of cfDNA for the diagnosis of NSCLC found a pooled sensitivity of 68% via a systematic review^18^. Currently, the NCCN guidelines only endorse 1) a plasma-first approach for testing for EGFR T790M in patients who have developed resistance to first- or second-generation tyrosine kinase inhibitors (TKIs), with tissue biopsy being recommended in cases where plasma testing is negative^8^, and 2) liquid biopsy in specific clinical circumstances where the patient is medically unfit for invasive tissue sampling or when tumor tissue specimen is inadequate or unobtainable, following pathological confirmation of diagnosis, with a follow-up tissue-based analysis in cases where no oncogenic driver is identified in plasma cfDNA^8^. This is aligned with the latest recommendations from the International Association for the Study of Lung Cancer (IASLC) for liquid biopsy for NSCLC, where liquid biopsy is recommended for cases where tissue sample is unavailable (‘plasma first’ approach), or in cases where tissue biopsy is inadequate for conducting comprehensive tissue genotyping (‘complementary’ approach)^19^. Furthermore, according to the IASLC recommendations, for cases with oncogene-addicted NSCLC progressing after initial targeted therapy, a ‘plasma first’ approach should be considered standard of care^19^.

Liquid and tissue biopsies each present their own strengths. In this study, we hypothesize that plasma cfDNA testing using a panel of target genes can complement standard molecular testing using tissue biopsy for NSCLC patients. Here, standard molecular tests encompass single target (e.g. *EGFR*) PCR-based tests, which could require time-consuming serial tissue testing depending on previous findings. For plasma cfDNA testing, next-generation sequencing (NGS)-based approaches, if adequately sensitive and comprehensive, have been shown to identify actionable mutations in plasma cfDNA of advanced NSCLC^20,21^. Therefore, rather than substituting tissue biopsies with liquid biopsies, adding a concurrent plasma NGS test to tissue testing would improve the detection of actionable mutations in patients with NSCLC, improving prognostication in addition to choice and timeliness of treatment initiation. This may translate to a ‘plasma-first’ approach where getting a tissue sample is rendered impractical or extremely difficult^19^.

This study focuses on standard tissue testing for mutations in the *EGFR* gene, which is mutated in 40-60% of Asian patients, and 10-20% of Caucasian NSCLC patients^22^. Specifically, EGFR L858R and in-frame exon 19 deletions, account for 50% and 40% of *EGFR* mutations, respectively, and are sensitizing mutations as tumors harboring these mutations are sensitive to EGFR tyrosine kinase inhibitors (TKI)^23^. Molecular testing for alterations in multiple genes such as *EGFR, ALK, ROS1, RET, BRAF, ERRB2, MET* exon 14 and NTRK1/2/3 have progressively entered the standard of care over the last 10 years ^24^. Here, we aim to demonstrate the clinical utility of an ultrasensitive, amplicon-based NGS tool for plasma cfDNA testing alongside standard tissue testing in patients suspected to have lung cancer, for the purposes of widening the scope of eligibility for treatment and reducing waiting time for molecular test results.

## METHODS

### Study design and patients

Patients with suspected lung cancer were prospectively enrolled for this study at the Department of Respiratory Medicine, Changi General Hospital, Singapore between June 2015 to August 2018. Prior to diagnosis by histology, blood samples for NGS-based plasma genotyping were collected during patient visit, followed by baseline tissue sampling by bronchoscopy or effusion collection within 48 hours. Patients were subsequently diagnosed to have non-small cell lung carcinoma (NSCLC), other cancers, or not cancer based on histology, cytology or microbiological testing. For NSCLC patients, standard of care targeted *EGFR* mutation tissue testing was performed on tumor biopsy samples, where available, using the Roche cobas® EGFR Mutation Test or by Sanger sequencing in a College of American Pathologists (CAP)-accredited clinical laboratory, and results were available as clinical reports. For all patients with blood available, targeted NGS plasma testing was performed in a CAP-accredited clinical laboratory, to detect tumor mutations in cfDNA. Similar targeted NGS testing was also performed in matched tissue samples, for cases with additional tissue available. Basic patient characteristics, including age, gender and confirmed histological diagnosis were recorded as part of the study. This study was approved by the institutional review board of Changi General Hospital and is registered under clinical trial number NCT04254497.

### Plasma and Tissue NGS Genotyping

Peripheral blood was collected in EDTA tubes and blood was processed within 24 hours of collection to isolate plasma. Circulating nucleic acid was extracted from plasma samples using the QIAamp Circulating Nucleic Acid Kit (Qiagen) and cfDNA was used to perform a next-generation sequencing (NGS) assay (LiquidHALLMARK®) in a CAP-accredited clinical laboratory. LiquidHALLMARK® is a clinically validated, ultrasensitive, amplicon-based assay for the detection of single nucleotide variants (SNVs), insertion-deletion mutations (indels) and copy number alterations among 49 genes (at the time of this study) (**Table S1**) with sensitive detection at variant allele frequencies above 0.1% for SNVs and indels. In this study, clinically actionable mutations were defined as mutations in *EGFR, ERBB2, BRAF, KRAS*, and *MET* (exon 14 skipping and copy number gains) which are therapeutically targetable, guideline recommended biomarkers or emerging biomarkers for the treatment of metastatic NSCLC^25^. Tumor DNA was extracted from remaining available tissue biopsy material using the QIAamp DNA FFPE Tissue Kit (Qiagen) and was also analyzed for panel-wide confirmation and concordance of findings from plasma cfDNA, using the same platform technology as LiquidHALLMARK® (TissueHALLMARK®). The NGS assay did not examine fusions in *ALK, RET* and *ROS1* at the time of this study.

### Data Analysis

Concordance analysis between routine molecular tissue testing and plasma samples was focused on the presence of mutations in *EGFR* as this is a routine molecular diagnostic test available for NSCLC patients, ordered by practicing oncologists. Sensitivity and specificity analyses were performed taking tissue *EGFR* test results as standard. Other actionable mutations (non-*EGFR*) detected in *BRAF, KRAS, ERBB2* and *MET* using plasma NGS panel testing were recorded as additional actionable findings, and any other mutations detected among the 49 genes targeted in the NGS assay were recorded as other genomic findings. The overall rate of detection of mutations in plasma cfDNA NGS was analyzed. For panel-wide testing done on matched plasma and tissue samples (where available) positive and negative predictive agreement analysis was performed for all actionable genomic findings. For NGS, variant allele frequencies (VAFs) were analyzed and are defined as the proportions of variant alleles relative to wild-type alleles. For patients with concurrent plasma and tissue NGS tests, correlation analysis of plasma and tissue variant allele frequencies (AFs) was done using Spearman rank correlation. Fisher’s exact test was used to determine associations between detection of actionable mutations and average coverage, and disease stage for NSCLC. All analyses were performed using RStudio V1.2.5033.

Clinical endpoints included test turnaround time (TAT), measured in days from biopsy sampling to reporting of *EGFR* molecular test results, or from blood sampling to reporting of NGS results.

## RESULTS

### Patient and Sample Characteristics and Test Results Accessibility

A total of 71 patients suspected to have lung cancer, based on their diagnostic scans and symptomatology, were enrolled. Patients were predominantly male (52/71, 73%) and the median age of the patient group was 67 years (range 31-87). Based on histology or cytology specimens, 54 patients (76.1%) were subsequently confirmed to have NSCLC, 7 (9.9%) were diagnosed with having other cancers, and the remaining 10 (14.1%) did not have cancer, with diagnosis of tuberculosis, pneumonia or inflammation, or an undetermined non-cancer diagnosis (**Table 1**). Blood samples could be obtained for 99% (70/71) patients prior to diagnostic biopsy sampling. Among NSCLC cases, blood sample was not available for 1 patient, resulting in an accessibility rate of 98.4% (53/54) for blood samples. Plasma NGS testing was successful for 100% of blood samples collected (70/70), and among 100% of NSCLC patients with blood samples available (53/53). Volume of plasma available ranged from 0.5 to 9 ml (median, 5.5 ml), and yield of cfDNA was in the range of 10-350 ng per ml plasma (median, 19.24 ng per ml plasma) (**Figure S1**).

**Table 1.**
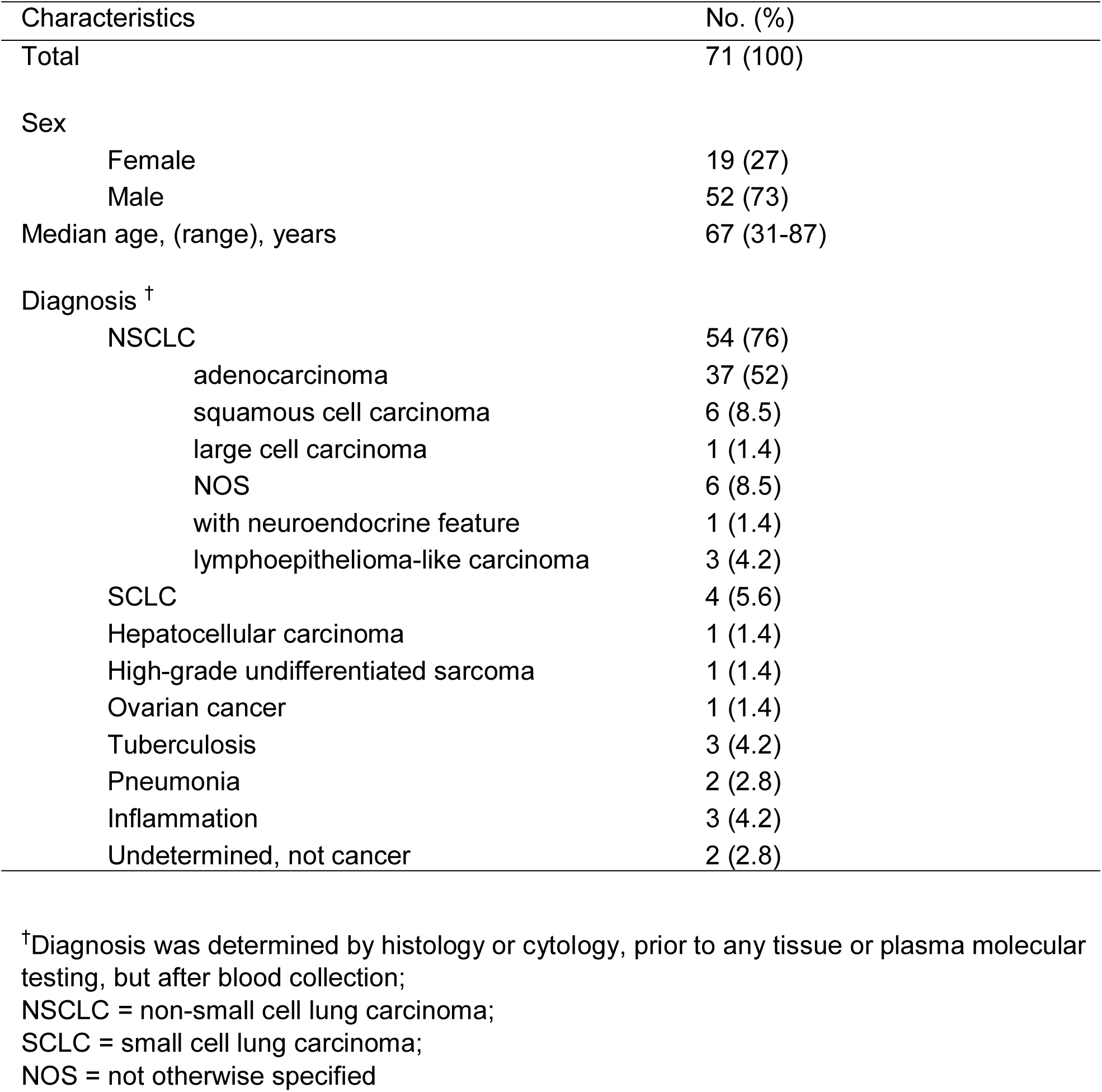
Baseline Patient Characteristics.

| Characteristics | No. (%) |
| --- | --- |
| Total | 71 (100) |
| Sex |  |
| Female | 19 (27) |
| Male | 52 (73) |
| Median age, (range), years | 67 (31-87) |
| Diagnosis <sup>†</sup> |  |
| NSCLC | 54 (76) |
| adenocarcinoma | 37 (52) |
| squamous cell carcinoma | 6 (8.5) |
| large cell carcinoma | 1 (1.4) |
| NOS | 6 (8.5) |
| with neuroendocrine feature | 1 (1.4) |
| lymphoepithelioma-like carcinoma | 3 (4.2) |
| SCLC | 4 (5.6) |
| Hepatocellular carcinoma | 1 (1.4) |
| High-grade undifferentiated sarcoma | 1 (1.4) |
| Ovarian cancer | 1 (1.4) |
| Tuberculosis | 3 (4.2) |
| Pneumonia | 2 (2.8) |
| Inflammation | 3 (4.2) |
| Undetermined, not cancer | 2 (2.8) |

<sup>†</sup>Diagnosis was determined by histology or cytology, prior to any tissue or plasma molecular testing, but after blood collection;
NSCLC = non-small cell lung carcinoma;
SCLC = small cell lung carcinoma;
NOS = not otherwise specified

Patients with NSCLC (n = 54) were eligible for tissue *EGFR* testing, however, tissue *EGFR* test results were available only for 38 NSCLC patients, resulting in a significantly lower tissue results accessibility rate of 70.4% (38/54), with 29.6% of cases (16/54) having no *EGFR* test results from tissue (**Figure S2**). There were two primary reasons for lack of tissue *EGFR* test results for NSCLC patients, including failure to obtain adequate biopsy sample due to patients’ advanced age or aggressive disease in 37.5% of cases (6/16), and failure to obtain informative *EGFR* test results from collected biopsies for 62.5% of cases (10/16). Patient enrollment, testing workflow and an overview of mutation findings are described in **Figure 1**.

**Figure 1.**
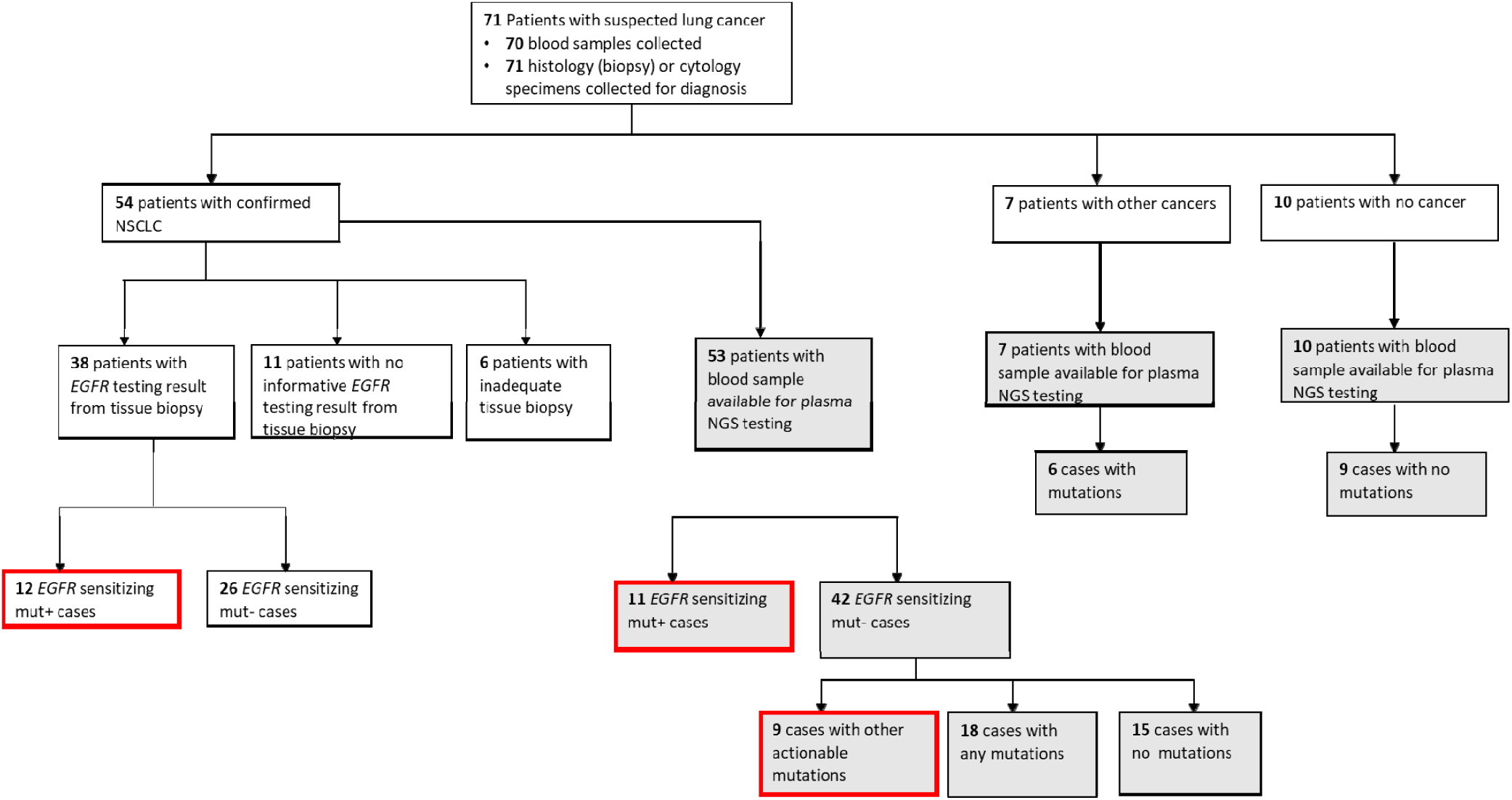
Patient enrolment and testing workflow. Flowchart showing the patient enrolment, diagnosis, sample availability, type of testing conducted an mutation findings. Grey-filled boxes indicate testing results from plasma NGS, and red outlined boxes are all clinically actionable findings made in this stud for NSCLC cases.

Among NSCLC cases with successful tissue *EGFR* testing using biopsy, an *EGFR* mutation was found in 31.6% of cases (12/38), whereas an *EGFR* sensitizing mutation was found in 20.7% of cases (11/53) that underwent plasma NGS testing. Among NSCLC cases tested by plasma NGS that were negative for *EGFR* sensitizing mutations, additional actionable findings were made in 9 of 42 cases (21.4%), and other genomic findings (any other non-actionable mutations from the 49 gene LiquidHALLMARK® panel) were made in 42.8% (18/42) cases. Of the 7 cases subsequently diagnosed by histology or cytology to have other non-NSCLC cancers, 6 cases had ≥1 mutation identified by plasma NGS testing. In 9 of 10 patients with non-cancer diagnosis confirmed, no mutations were detected by plasma NGS testing.

### Diagnostic Yield from Tissue Biopsy and Plasma

Diagnostic yield was compared for NSCLC patients where testing was possible with either standard tissue *EGFR* test with concurrent plasma NGS testing, or with plasma NGS only, as dictated by sample availability (**Figure 2A**). Of the NSCLC cases with available tissue *EGFR* test results, 31.6% (12/38) were positive for *EGFR* sensitizing mutations while the remaining 68.4% (26/38) had a negative *EGFR* mutation finding. Of these 38 cases, 37 cases were also tested by plasma NGS (blood was not available for one case) with 24.3% (9/37) having a positive result for *EGFR* sensitizing mutation and the remaining 75.7% (28/37) having a negative *EGFR* mutation result (**Figure 2B**). Specifically, among tissue *EGFR*-negative cases also tested by plasma NGS (n = 25), plasma NGS did not identify any further *EGFR* sensitizing mutations (for which FDA-approved therapies are available) but did identify other clinically actionable mutations in 6 cases, including *MET* exon 14 skipping (n = 1), BRAF V600E (n = 1), BRAF K601E (n = 1), KRAS G12D (n = 2), EGFR exon 20 insertion (n = 1) (**Figure 2B**).

**Figure 2.**
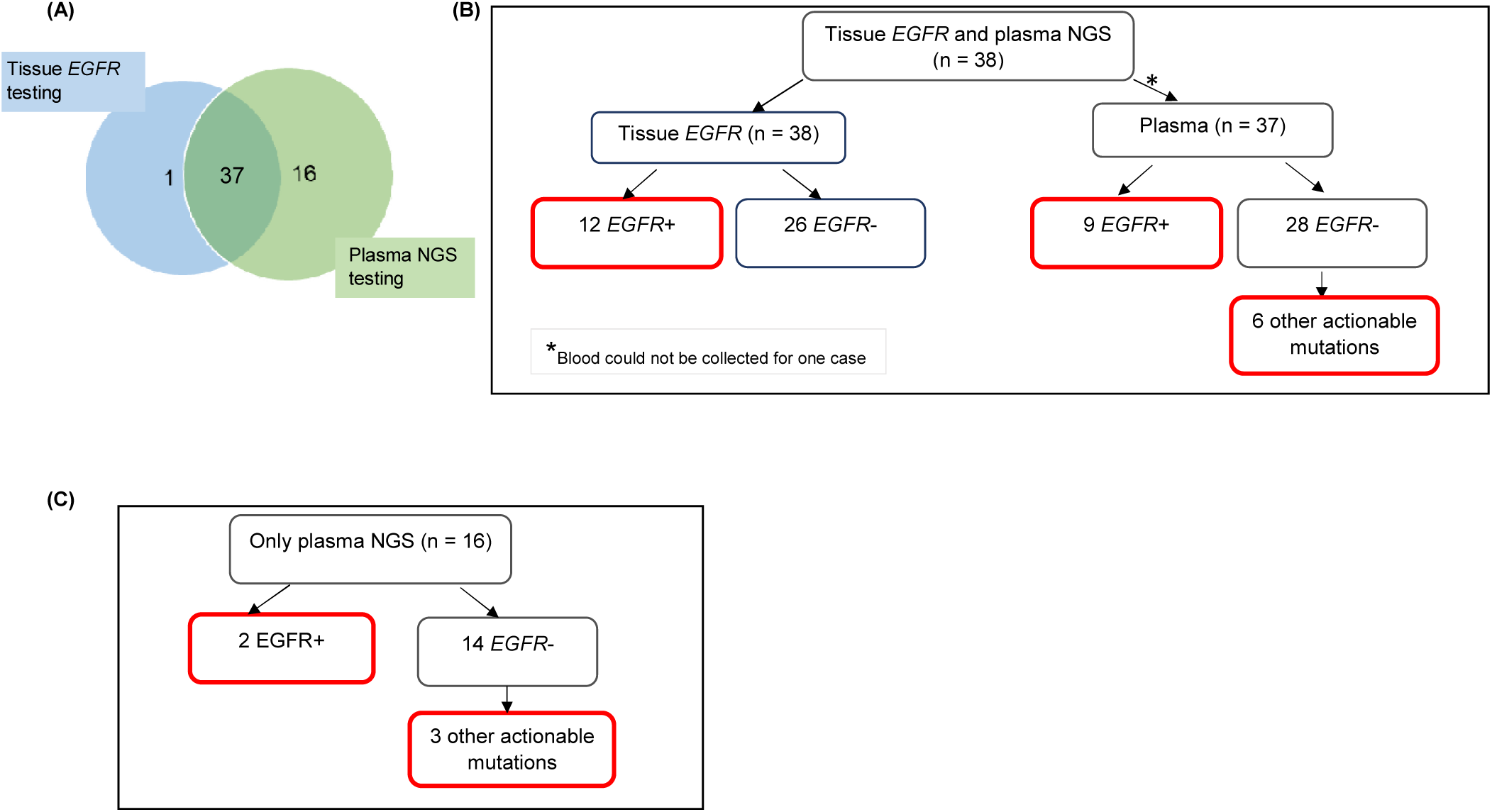
Diagnostic yield from molecular testing of tissue and plasma samples for 54 NSCLC patients. **(A)** Nearly all 38 patients with informative tissue *EGFR* testing underwent plasma NGS testing (except one). An additional 16 patients had only plasma NGS testing done, due to inadequate tissue biopsy sample for molecular testing or non-informative results from tissue testing. **(B)** Findings of *EGFR* sensitizing mutations and other actionable mutations in cases with both tissue *EGFR* and plasma NGS results. Six other actionable mutations from plasma NGS testing included *MET* exon 14 skipping (n = 1), BRAF V600E (n = 1), BRAF K601E (n = 1), KRAS G12D (n = 2), EGFR exon 20 insertion (n = 1) **(C)** Clinically actionable findings in cases with only plasma NGS testing. Boxes outlined in red indicate clinically actionable diagnostic yield from all testing modalities.

Importantly, where tissue *EGFR* testing results were lacking and only plasma NGS was performed (n = 16), clinically actionable mutations were detectable in 5 cases, including sensitizing *EGFR* mutations E746_A750del (n = 1) and L747_P753delinsS (n = 1), BRAF K601E (n = 1), KRAS G12D (n = 1) and *MET* exon 14 skipping (n = 1) **(Figure 2C)**.

The additional diagnostic yield from plasma NGS testing for tissue *EGFR*-negative cases is therefore 24% (6/25), for which other actionable mutations were detected. Among NSCLC samples which totally failed to undergo tissue *EGFR* testing (n = 16), plasma NGS provided a diagnostic yield of 31.3% (5/16). The total additional diagnostic yield by plasma NGS, is therefore 26.8% (11/41).

In this cohort of 54 NSCLC patients, irrespective of availability of tissue *EGFR* testing, a plasma NGS test on its own would have provided clinically actionable mutation information in up to 37% of cases (20/54). In contrast, standard tissue *EGFR* testing (with limitations of tissue sampling and quality and breadth of testing), accurately identified only 22.2% (12/54) of cases with clinical actionability based on *EGFR* sensitizing mutations. Performing both tissue and plasma testing resulted in a diagnostic finding in 42.6% (23/54) of NSCLC cases, considering only tissue *EGFR* test and plasma NGS test not including *ALK, RET, ROS1* fusions among actionable targets, which represents a 1.9-fold increase in the number of actionable findings compared to tissue *EGFR* testing alone.

The spectrum of all mutations (actionable and non-actionable) detected by plasma NGS in 53 NSCLC cases is shown in **Figure 3**. A total of 38 NSCLC cases (76%) had ≥1 alteration detectable, of which *TP53* mutations were most prevalent (60.5%), followed by mutations in clinically actionable target genes, *EGFR* (31.6%), *BRAF* (13.2%), *KRAS* (10.5%), and *MET* (7.9%).

**Figure 3.**
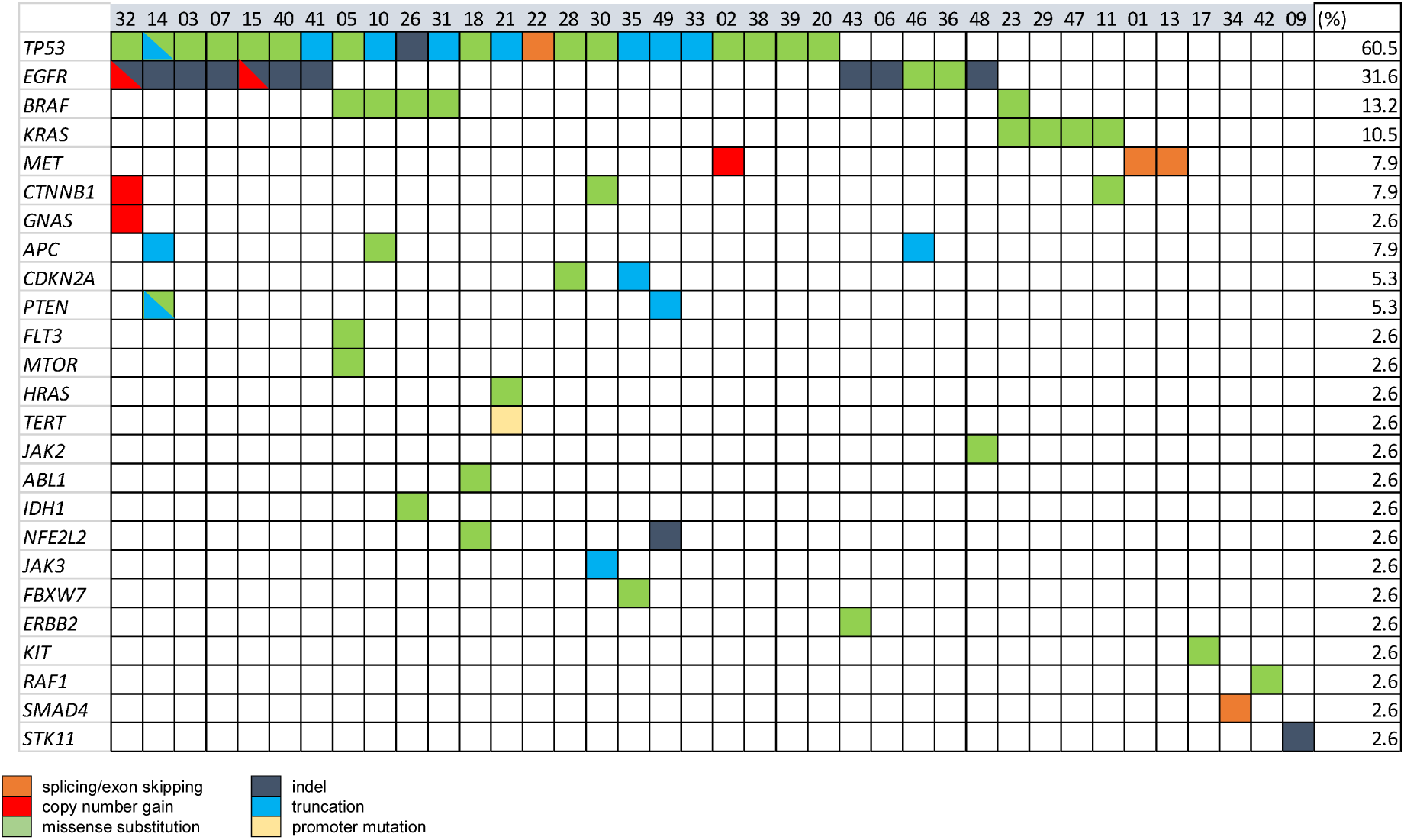
Spectrum of genomic alterations in NSCLC detected by plasma NGS testing. Baseline plasma samples for 53 NSCLC cases were tested 49-gene panel LiquidHALLMARK® assay. Cases with ≥1 alteration are presented (n = 38), 15 cases with no alteration detected were excluded from presentation. Genes with no alteration detected among all cases were also excluded from presentation. Percentage of cases carrying a mutation in each gene are shown on the right-most column (%).

### Tissue and Plasma Concordance for EGFR mutations and other Variants

In order to assess the performance of plasma NGS test relative to the standard tissue *EGFR* testing modality, samples with results from both plasma NGS and tissue tests were compared. Among 12 cases positive for sensitizing *EGFR* mutations by standard *EGFR* tissue testing, 9 were found to have the same mutation in plasma cfDNA (**Figure 4A**), for a sensitivity of 75% (9/12). Out of 26 cases negative for sensitizing *EGFR* mutations in tissue, 25 cases were tested by plasma NGS, and concordantly none were found to have any *EGFR* mutations (except for one case with an *EGFR* exon 20 insertion (EGFR A763_Y764insFQQA)) resulting in a specificity of 100% (25/25). The overall concordance of *EGFR* sensitizing mutations commonly included in the range of PCR-based *EGFR* testing and plasma NGS was 91.9% (34/37). The range of *EGFR* VAFs detected by plasma NGS was 0.057-80.3%, with a median AF of 0.98%, with 8 *EGFR* exon19 deletions and 2 L858R mutations (**Figure 4B**). As the detection sensitivity of NGS assays is a function of the depth of coverage achieved, which in turn is a function of input DNA amount, we looked at the distribution of depth of coverage across the samples for which *EGFR* mutations were expected to be found in plasma based on tissue results. For three samples in which corresponding *EGFR* mutations were not detected in plasma, the average consensus coverage (X) was 6524x, 8068x and 14565x, respectively (**Figure 4B**), which did not correspond to the lowest coverage among these samples. In fact, two cases with coverage of 4538x and 4830x, respectively had detectable mutations at variant allele frequencies of 0.057% and 9.44% for EGFR E746_A750del (exon 19 deletion), suggesting a biological (such as low tumor shedding into circulation) rather than a technical reason for discordance. Considering all samples tested by plasma NGS (n = 53), the median consensus coverage was 8183x. Among samples with coverage lower than the median coverage (n = 26), 9 samples had no mutations detected by plasma NGS, and among samples with coverage greater than or equal to the median coverage (n=27), 6 samples had no mutations detected by plasma NGS (p=0.2238, Fisher’s exact test), suggesting that coverage was not the main determining factor for detection of variants among these samples (**Figure S3**).

**Figure 4.**
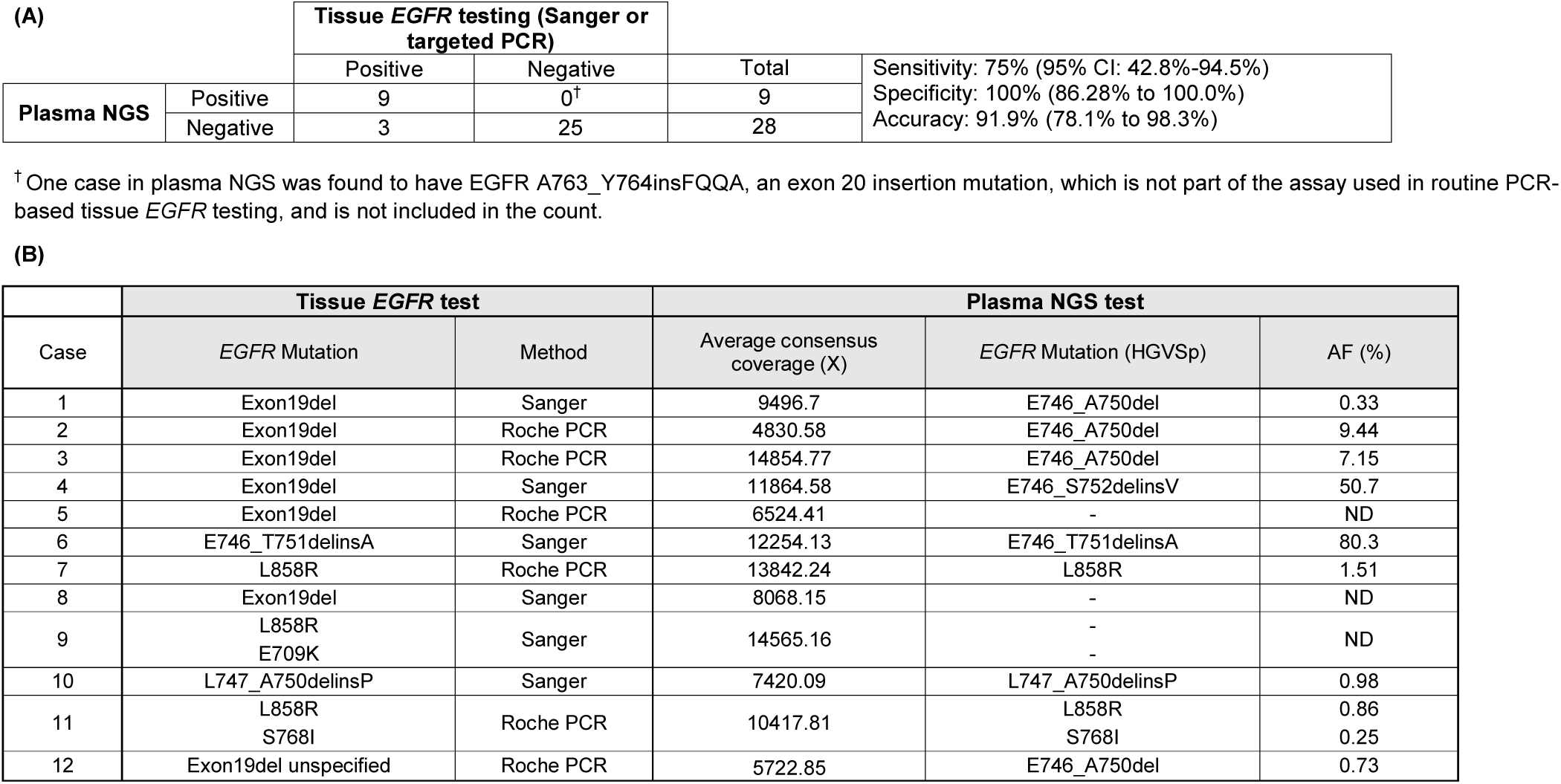
Concordance analysis of *EGFR* mutation detection by targeted tissue *EGFR* testing and plasma NGS for 37 NSCLC cases. **(A)** Sensitivity of plasma NGS for *EGFR* detection relative to tissue *EGFR* testing was 75% (9/12) and specificity was 100% (25/25), for an overall concordance of 91.9% (34/37). **(B)** Depth of coverage by plasma NGS and detection of *EGFR* mutation and mutation allele frequency (AF %) are not directly related. ND, not detected.

Beyond *EGFR*, panel-wide concordance of mutation findings in tissue biopsy samples and plasma was studied by performing tissue NGS using the same panel (TissueHALLMARK) on a subset of samples for which tissue samples from original biopsy were available. A total of 24 NSCLC patients had both plasma NGS and tissue NGS results available, of which 14 (58.3%) cases had a therapeutically relevant target detected, either by plasma or tissue NGS or by both methods. The positive predictive agreement (PPA) between plasma and tissue NGS was 75.0% and the negative predictive agreement (NPA) was 83.3%, for an overall predictive agreement (OPA) of 79.2% (**Figure 5A)**. There was a correlation between the plasma and tissue mutation AF among actionable mutations detected (ρ = 0.5503, p-value = 0.0221) (**Figure S4**). It was observed that for cases in which tissue mutation was not detected in plasma, the AF was low in the tissue sample, below 10% AF. Conversely, two mutations identified only in plasma were characterized by very low AF - 0.04% and 0.3% **(Figure S4)**. To account for the discordant mutations, the extent of clinical disease was examined by comparing tumor stage information, which was available only for 49% of NSCLC cases in this study that underwent plasma NGS (26/53). Actionable mutations were detected in plasma for 0% (0/6) cases with disease stage 2B-3B, including one tissue-discordant *EGFR* sensitizing mutation (**Figure 5B**). In contrast, for cases with disease stage 4 or 4B (n = 20), an actionable mutation was detected in 45% (9/20) cases, including 5 tissue concordant *EGFR* sensitizing mutations (Stage 2B-3B vs Stage 4-4B: Fisher’s exact test, p-value = 0.0632) (**Figures 5B and 5C**).

**Figure 5.**
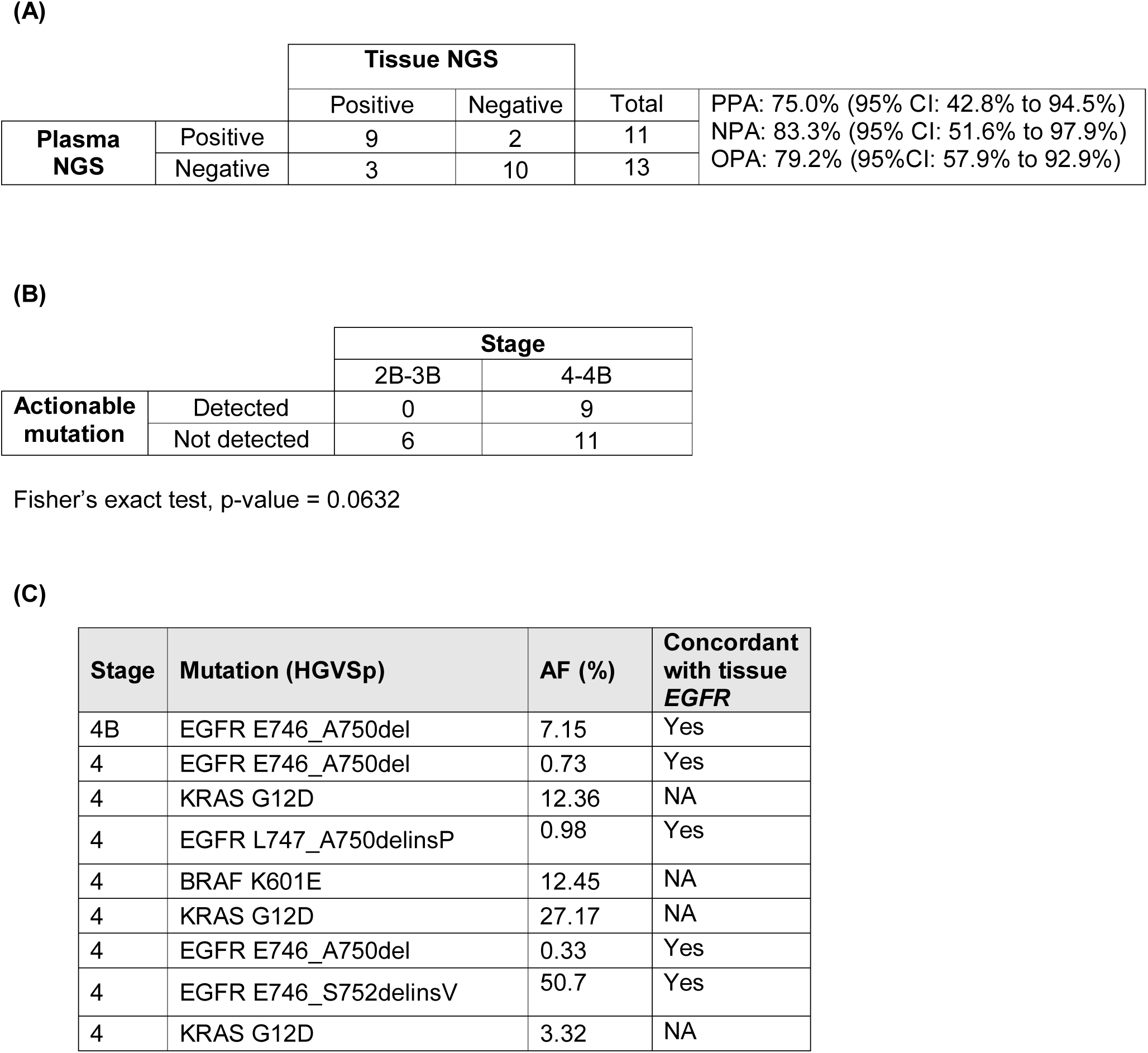
Analysis of actionable mutation detection by plasma NGS for NSCLC cases. **(A)** Panel-wide concordance of actionable mutations in 24 NSCLC cases that underwent both tissue and plasma NGS testing. **(B)** Detection of actionable mutations in plasma was cancer stage-dependent, with 9 (of 20) stage 4-4B NSCLC cases with detectable actionable mutations, compared to 0 (of 6) stage 2B-3B NSCLC, Fisher’s exact test, p-value = 0.0632. **(C)** Clinically actionable mutations detected in stage 4-4B cases and their concordance of detection with tissue *EGFR* tests. NA = not applicable as not tested by tissue *EGFR*.

### Plasma NGS for non-NSCLC cancers and non-cancer samples

As described in **Figure 1**, plasma samples from patients initially suspected to have lung cancer, but later confirmed to have either other cancers (n = 7) or a non-cancer diagnosis (n = 10), were also tested by NGS. The specificity of detection of cancer-specific mutations by plasma NGS was demonstrated by the detection of a mutation in 85% (6/7) of other cancer cases, including pathogenic *TP53* mutations in 71% (5/7) of cases (**Figure S5**). Importantly, among plasma from 10 non-cancer cases, only one case harboured an *ALK* frameshift mutation of uncertain significance, which was also present in a pleural effusion sample from the same case (not shown). This demonstrates that mutation detection by plasma NGS is reliable and specific to the presence of cancer.

### Plasma NGS turnaround time (TAT)

Plasma NGS was successfully performed in 53 NSCLC patients and 17 non-NSCLC patients with suspected lung cancer with an average TAT of 10 days from the time of blood draw to the time of receipt of report. In contrast, the average TAT for tissue NGS for 38 patients with standard *EGFR* testing with tissue was 29.9 days (p-value <0.05), with the longest duration between biopsy collection and receipt being 48 days.

## DISCUSSION

In this single-centre prospective study we assessed the clinical utility of adding plasma NGS testing to the diagnostic workflow for suspected lung cancer and molecular testing workflow for diagnosed NSCLC. This approach may be labelled as ‘plasma-first’ for cases with no tissue sample available for testing, or complementary where both tumor and plasma sample may be tested for comprehensive target coverage, or where there is uncertainty about adequacy of tissue sample for molecular testing^19^. Plasma NGS demonstrated significantly higher sample accessibility levels, lower average reporting time and matched specificity and accuracy when compared to standard tissue *EGFR* testing. Importantly, a range of additional actionable mutations from guideline-recommended biomarkers were found by plasma NGS, potentially enabling an appropriate targeted treatment option, even in the absence of a tissue test result.

The invasive nature of tissue biopsy makes routine diagnostic EGFR profiling unfeasible for patients with late-stage NSCLC and those of advanced age. In this study, only 70.3% (38/54) of diagnosed NSCLC patients had informative results from tissue *EGFR* testing. On the other hand, blood sample was collected for 99% of all patients recruited in this prospective study (70/71), including 98.4% (53/54) of NSCLC patients. We show the clinical value of a plasma NGS test was an average 26.8% additional diagnostic yield over tissue *EGFR* testing, from the combined contribution of 1) additional actionable mutations in 6 of 25 tissue *EGFR*-negative cases and 2) detection of 5 actionable mutations, including 2 *EGFR* sensitizing mutations, in 16 cases that had no results from tissue *EGFR* testing.

Among all NSCLC patients, adding plasma NGS to tissue *EGFR* testing resulted in the detection of a therapeutically actionable mutation in 43.6% (23/54) cases, whereas if only tissue *EGFR* testing had been done, only 22.2% (12/54) cases would have had a clinical actionable finding. This represents a 1.9-fold increase in the number of actionable mutations detected in this study by adding plasma NGS testing, including two *EGFR* mutations in two cases which failed standard tissue EGFR testing. This is consistent with past studies in larger real-world NSCLC cohorts, where the addition of comprehensive liquid biopsy to targeted tissue testing increased the number of targetable mutations up to as much as 65%^17,26^. This makes plasma NGS an especially important diagnostic tool when tissue biopsy is scant or not available.

The routine implementation of a complementary plasma or even ‘plasma-first’ testing approach in healthcare settings significantly reduces reporting time and can enable patients to begin targeted therapy earlier. Tissue *EGFR* results took an average of 29.9 days to report, while plasma NGS took an average of 10 days to report. In this study, for 37% (20/54) of NSCLC cases, a treatment decision could have been made as soon as the plasma NGS results became available. This trend of a lower turnaround time with plasma NGS tests has been widely supported by other studies^27,28^. The length of time between the scheduling of the tissue biopsy and the procedure itself can vary widely and can add many weeks to an already long wait for a diagnosis. In contrast, in-clinic, same-day blood collection for plasma NGS can be quickly and conveniently performed.

High specificity of diagnostics is key to ensuring that false positive findings do not result in incorrect treatment, which can be harmful to patients and increases the financial burden of healthcare. The specificity of plasma NGS compared with PCR-based tissue *EGFR* testing in this study was 100%. This finding provides supporting evidence that positive identification of an actionable mutation by plasma NGS is sufficient evidence to initiate targeted treatments without needing additional confirmation from tissue testing^26^, reducing the duration from clinical consultation to the start of the treatment program. In this study, tissue *EGFR* results would have yielded additional findings in 5.56% (3/54) of cases, for which plasma NGS did not find the *EGFR* mutation present in tumor, supporting the complementary plasma testing would be the most informative approach for the NSCLC patient population.

Plasma NGS reported a sensitivity of 75% when compared to routine Sanger or targeted PCR, which suggests that negative results require further investigation to rule out the possibility of false negatives. Levels of circulating tumor DNA (ctDNA) are highly varied between patients, likely because ctDNA levels can vary based on the rate of turnover, perfusion and vascularisation of the tumor, and are influenced by the cancer stage^29^. In this study, a disease stage-dependence was observed for both *EGFR* mutation concordance as well as detection rate of any actionable mutation, with 45% (9/20) of cases with stage 4 or 4B versus 0% (0/6) of cases with stage 2B-3B having an actionable mutation detected. Further, there was a correlation between the tumor AF and plasma AF of mutations for cases where both plasma and tissue NGS was performed, suggesting ctDNA burden is a function of the actual tumor size and spread. This is in alignment with another study where patients with liver metastases had higher plasma-tissue concordance for actionable mutations compared to those with M1a disease^17^, and with a study in which patients with intrathoracic metastases alone were less likely to have detectable ctDNA^30^. It has been suggested that disease stage could serve as a decision metric to decide the order in which plasma or tissue testing is requested, to maximize detection of actionable mutations detection without unnecessarily prolonging the time to result.

As an attestation of the specificity and broad applicability of plasma NGS for cancer diagnostics, we also show that among other suspected lung cancer patients eventually diagnosed to not have cancer, only one case (out of 10) had a detectable mutation (of uncertain significance), while 6 of 7 cases diagnosed as having other non-NSCLC cancers had *TP53* mutations or other cancer-related mutations detected. Based on these results, a role for plasma NGS testing in preliminary cancer diagnosis could be envisioned.

This study has limitations in that the standard diagnostic test comparison was limited to *EGFR* mutations, and actionable fusions in *ALK, ROS1, RET* and *NTRK* were not considered as they were not measured at the time of this study. Including actionable fusions in concurrent tissue and plasma NGS tests will likely result in a similar fractional increase in actionability. Another limitation was that disease stage information was only available for a subset of patients, limiting the stage-specific analysis for plasma-tissue concordance. Further, the study was conducted in a small cohort of prospectively recruited patients and no information on treatment decisions and clinical outcomes was recorded, which would have enabled the real-world clinical utility of the complementary or ‘plasma-first’ approach to be quantified in a prospective setting. Finally, longitudinal monitoring of the efficacy of plasma NGS on this patient cohort was not captured in this study. However, the non-invasive nature and sensitive detection ability of plasma NGS make it a suitable tool for determination of resistance mutations earlier and with greater accessibility than would be possible with an initial biopsy or re-biopsy.

This study demonstrates that integrating plasma NGS with tissue testing increases actionable yield over conventional diagnostic approaches for NSCLC by allowing more patients to achieve comprehensive biomarker profiling. Plasma NGS allows for quick and non-invasive molecular profiling that can rapidly guide treatment decisions and complement routine tissue testing or tissue NGS, or could be a viable first-line alternative when tissue biopsy is not feasible.

## Data Availability

The datasets generated during and/or analysed during the current study are available from the corresponding author on reasonable request.

## ACKNOWLEDGEMENT

This study was funded by the Agency for Science, Technology and Research (A*STAR) Industrial Alignment Fund (IAF111221).

## Notes

### Competing Interest Statement

Yukti Choudhury, Min-Han Tan, Jonathan Poh and Kao Chin Ngeow are employees of Lucence Diagnostics

### Clinical Trial

NCT04254497

### Funding Statement

This study was funded by the Singapore Agency for Science, Technology and Research (A*STAR) Industrial Alignment Fund (IAF111221).

### Author Declarations

Changi General Hospital (Singapore) IRB.

