## Supplementary Data for "Complementing Tissue Testing with Plasma mutation Profiling Improves Therapeutic Decision Making for Lung Cancer Patients"

**Table S1. Composition of gene panel used for plasma NGS test (LiquidHALLMARK®).** The same gene panel was used for tissue NGS (TissueHALLMARK) for panel-wide comparison of mutations. *Genes for which copy number alterations can be calculated in both plasma and tissue tests.

| | LiquidHALLMARK® Panel (49 Genes) | | | | | | | | | |
|---|---|---|---|---|---|---|---|---|---|---|
| SNVs, indels | *ABL1* | *AKT1* | *ALK** | *APC* | *AR** | *ATM* | *BRAF* | *CCND1** | *CDH1* | *CDKN2A** |
| | *CTNNB1* | *EGFR** | *ERBB2** | *ESR1* | *FBXW7* | *FGFR2* | *FGFR3* | *FLT3* | *GATA3* | *GNA11* |
| | *GNAQ* | *GNAS* | *HNF1A* | *HRAS* | *IDH1* | *IDH2* | *JAK1* | *JAK2* | *JAK3* | *KIT* |
| | *KRAS* | *MAPK1* | *MAP2K1* | *MED12* | *MET** | *MTOR* | *MYC** | *NFE2L2* | *NOTCH1* | *NRAS* |
| | *PDGFRA* | *PIK3CA** | *PTEN** | *RAF1* | *SMAD4* | *STK11* | *TERT* | *TP53** | *VHL* | |

**Figure S1. Distribution of (A) volumes of plasma (ml) and (B) yields of cfDNA per ml of plasma for 70 patients.** Median and interquartile ranges are shown with red lines. **(C)** Yield of cfDNA per ml distribution by volume, dotted red line shows median cfDNA per ml (ng) amount = 19.24 ng.

**(A)** **(B)**

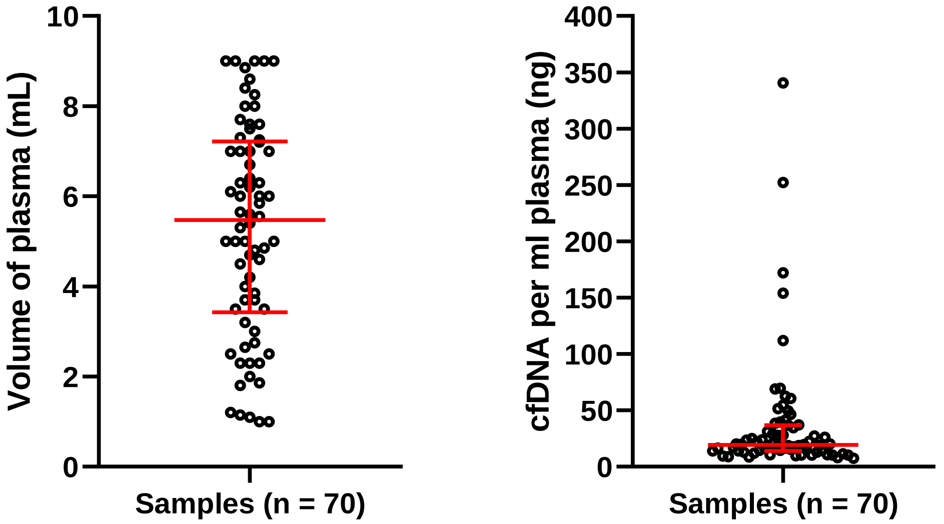


**(C)**

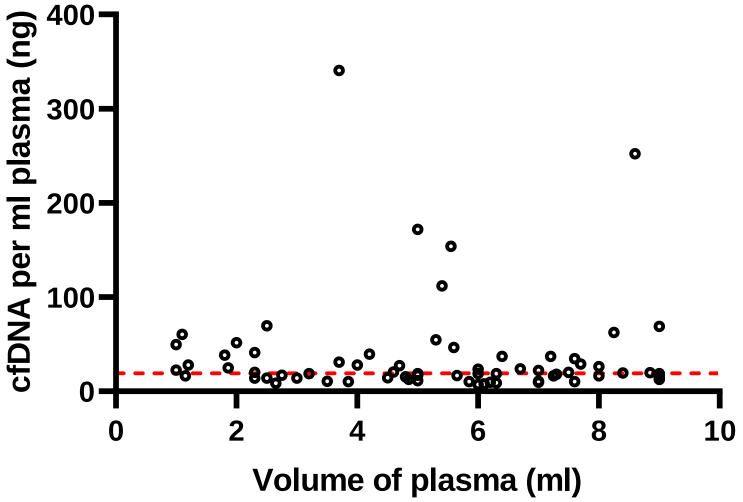

**Figure S2. Availability of *EGFR* test results from tissue biopsy samples among 54 NSCLC patients from a total 71 patients suspected to have lung cancer.**

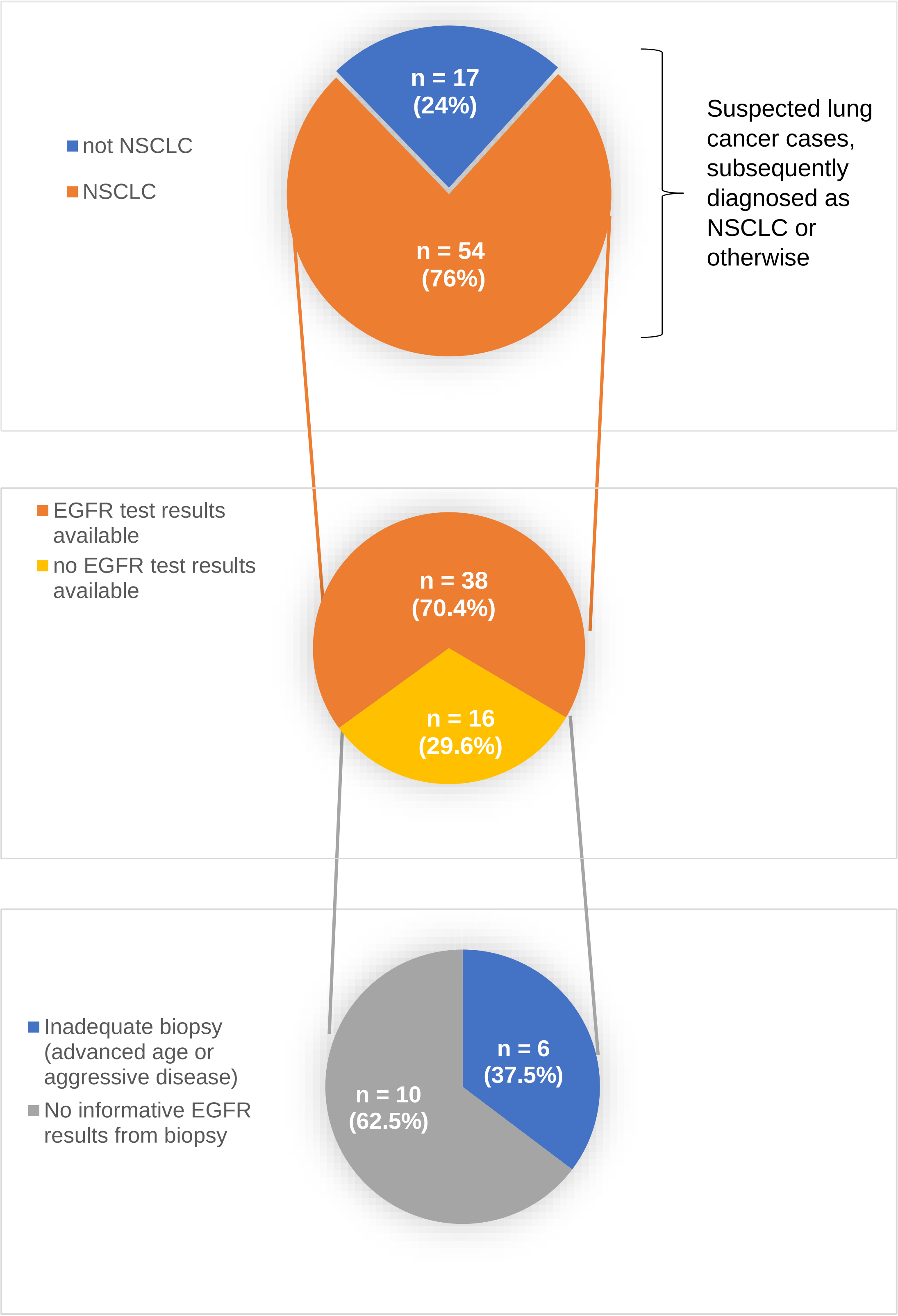

**Figure S3. Distribution of average consensus coverage for plasma NGS across 53 NSCLC samples. (A)** Average consensus coverage colored by detection of any mutations by plasma NGS, and samples in which *EGFR* mutation was not concordantly detected in plasma NGS. Median (8138x) and interquartile ranges for coverage are shown.

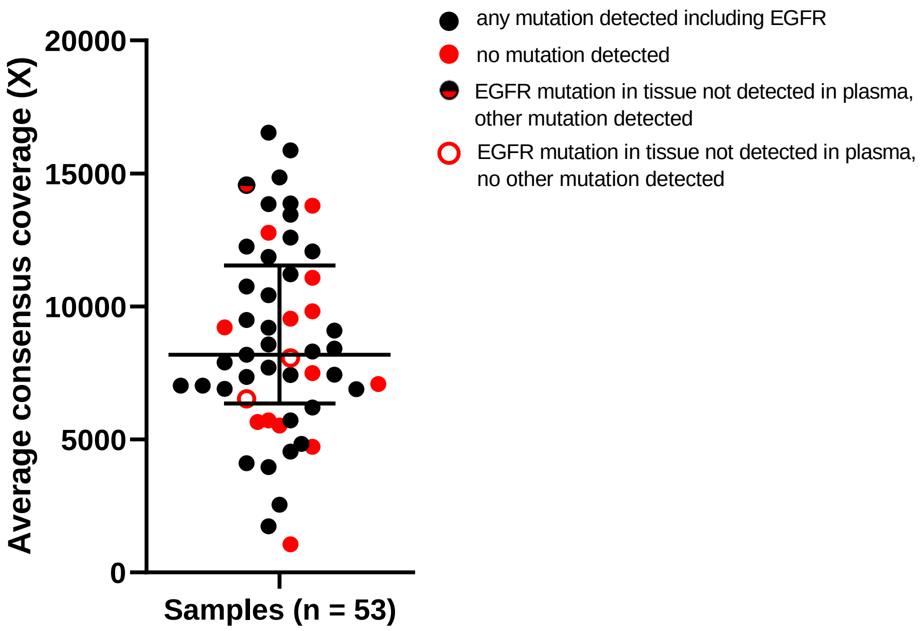

**Figure S4. Allele frequencies (AF) of therapeutically actionable mutations detected by plasma NGS and tissue NGS are correlated.** For the same patient's blood and tissue AFs of mutations detected are correlated (ρ = 0.5503, p-value = 0.0221). Red circles indicate cases where mutation was only detected in tissue, while green circles indicate mutations found only in plasma. All discordances were characterized by low detectable AFs, below 10% AF for mutations found only in tissue, and below 1% AF for plasma-only mutations.

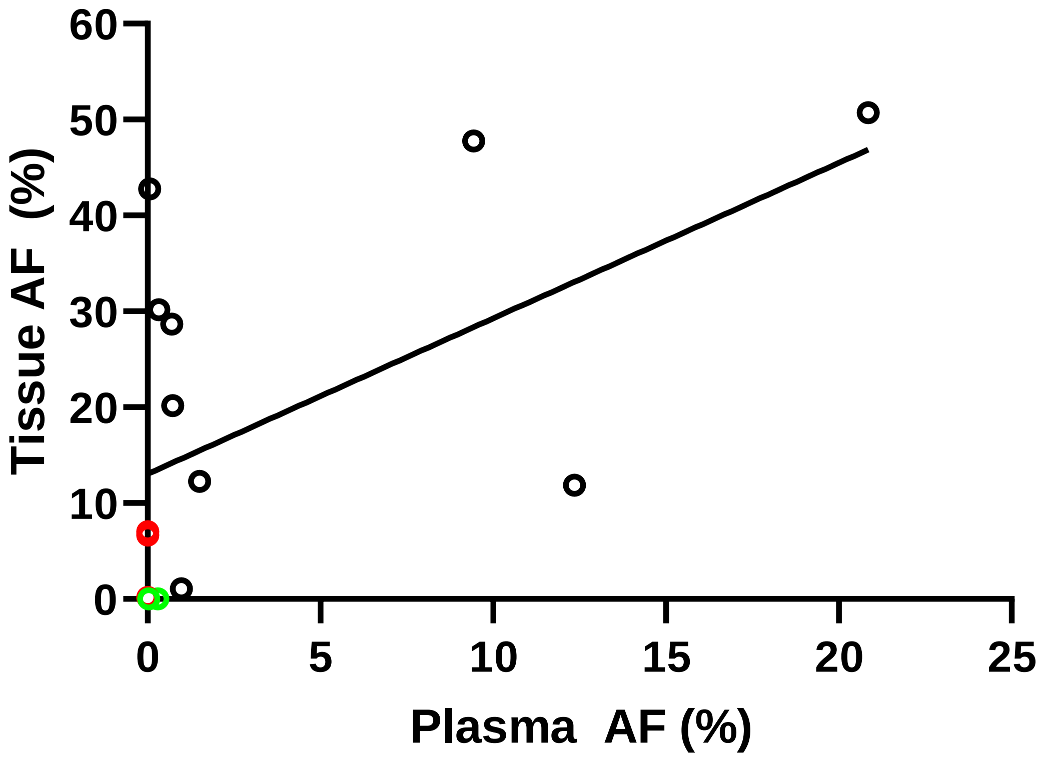

**Figure S5. Frequent detection of cancer-specific mutations for non-NSCLC cancer patients by plasma NGS.** SCLC = small cell lung carcinoma; HCC = hepatocellular carcinoma.

| Case | Diagnosis | Mutation (HGVSp) | AF (%) |
|---|---|---|---|
| 1 | SCLC | TP53 p.Pro278Arg | 27.11 |
| 2 | SCLC | TP53 p.Tyr163Cys | 12.78 |
| 3 | HCC | TP53 p.Arg249Ser<br>CTNNB1 p.Asp32Ala | 1.4<br>0.67 |
| 4 | High grade undifferentiated sarcoma | - | |
| 5 | Metastatic Ovarian Cancer | STK11 p.Pro315Leu | 0.38 |
| 6 | SCLC | TP53 p.Arg249GlyfsTer96 | 3.82 |
| 7 | SCLC | TP53 p.Gly154Val | 27.6 |
